# A phase 2 study of the inhaled pan-JAK inhibitor TD-0903 in severe COVID-19: Part 1

**DOI:** 10.1101/2021.03.09.21252944

**Authors:** Dave Singh, Maxim Bogus, Valentyn Moskalenko, Robert Lord, Edmund J. Moran, Glenn D. Crater, David L. Bourdet, Nathan D. Pfeifer, Jacky Woo, Elad Kaufman, David A. Lombardi, Emily Y. Weng, Tuan Nguyen, Ashley Woodcock, Brett Haumann, Rajeev Saggar

## Abstract

**Background:** Lung-targeted anti-inflammatory therapy could potentially improve outcomes in patients with COVID-19. The novel inhaled pan-Janus kinase (JAK) inhibitor TD-0903 was designed to optimise delivery to the lungs while limiting systemic exposure. Here, we report results from the completed Part 1 of a 2-part phase 2 trial (NCT04402866) in hospitalised patients with severe COVID-19.

**Methods:** Part 1 explored 3 doses of TD-0903 (1, 3, and 10 mg once-daily for 7 days) and placebo in a randomised, double-blind, ascending-dose study. Each dose cohort comprised 8 hospitalized patients (6:2 TD-0903:placebo) with PCR-confirmed COVID-19 requiring supplemental oxygen and receiving background standard-of-care therapy. Key objectives included safety and tolerability, pharmacokinetics, and oxygen saturation/fraction of inspired oxygen ratio; clinical outcomes were also explored. Data were summarised as descriptive statistics.

**Results:** Twenty-five patients were randomised to receive TD-0903 1 mg (n = 6), 3 mg (n = 7), 10 mg (n = 6), or placebo (n = 6). Almost all patients (92%) received background dexamethasone; 3 (12%) received remdesivir. TD-0903 was generally well tolerated with no drug-related serious adverse events. Low plasma concentrations of TD-0903 were observed at all doses. Clinically favourable numerical trends in patients receiving TD-0903 vs placebo included improved 8-point clinical status, shortened hospitalisation, improved oxygenation, and fewer deaths.

**Conclusions:** In Part 1 of this phase 2 trial, the novel inhaled JAK inhibitor TD-0903 showed potential for treatment of patients with severe COVID-19. TD-0903 3 mg is being evaluated in Part 2 of the randomised, double-blind, parallel-group trial in 198 hospitalized patients with COVID-19.

---

Severe coronavirus disease 2019 (COVID-19) is characterised by pneumonia with high levels of systemic inflammation, referred to as a “cytokine storm.”[1-3] Dexamethasone treatment decreases mortality in patients with COVID-19 who are receiving respiratory support and is now considered standard of care for patients with severe COVID-19.[4, 5] The orally administered Janus kinase (JAK)-1/2 inhibitor baricitinib in combination with the antiviral remdesivir has also shown clinical efficacy in patients with severe COVID-19.[6] JAK inhibition blocks cytokine signalling through signal transducer and activator of transcription (STAT) pathways, offering broad immunomodulation.[7] As pulmonary inflammation and associated diffuse alveolar damage drive COVID-19 morbidity and mortality,[3] lung-targeted anti-inflammatory therapy could potentially improve outcomes. The novel inhaled pan-JAK inhibitor TD-0903 was specifically designed to target all JAK isoforms (JAK1, JAK2, JAK3, TYK2) and optimise delivery to the lungs while limiting systemic exposure. Here, we report results from the completed Part 1 of a 2-part phase 2 trial (NCT04402866) in hospitalised patients with severe COVID-19.

This phase 2 study was designed with separate data reporting for Parts 1 and 2. Part 1 was a randomised, double-blind, placebo-controlled, multiple-ascending-dose trial conducted in the UK, Moldova, and Ukraine. The study protocol and related documents were approved by independent ethics committees at each site. The study was conducted in accordance with the principles of Good Clinical Practice and the Declaration of Helsinki. Patients provided informed consent. Patients 18 to 80 years of age with PCR-confirmed symptomatic COVID-19 (symptoms for 3–14 days) who were hospitalised and required supplemental oxygen to maintain saturation >90% were eligible. Patients receiving JAK inhibitors or anti–interleukin-6 therapy were excluded. Patients were sequentially enrolled in 3 ascending-dose cohorts (n = 8; 6 active and 2 placebo per cohort) and received once-daily TD-0903 1 mg (Day 1 loading dose 2 mg), 3 mg (Day 1 loading dose 6 mg), or 10 mg (no loading dose) or matched placebo via inhalation (Aerogen Solo + Ultra nebuliser system, Galway, Ireland) for up to 7 days, with follow-up through Day 28. Loading doses were administered on Day 1 for the 2 lowest maintenance dose levels (1 and 3 mg) to rapidly achieve pseudo-steady state in the lung.

Peripheral oxygen saturation (SaO2) was collected via pulse oximetry, and fraction of inspired oxygen (FiO2), vital signs, adverse events (AEs), and clinical status using an 8-point ordinal scale (OS)[8] were recorded daily though Day 7 and on Days 14, 21, and 28 and/or at hospital discharge. Physical examination, blood collection, and serum chemistry evaluations occurred on Days 1 and 7; patient care-related laboratory evaluations through Day 28 were also included. Key safety outcomes were change from baseline in vital signs and clinical laboratory results, and incidence and severity of treatment-emergent AEs (TEAEs; coded per Medical Dictionary for Regulatory Activities v23.1); key pharmacokinetic (PK) endpoints were plasma PK parameters on Days 1 and 7, and the key pharmacodynamic outcome was change from baseline SaO2/FiO2 ratio. Other clinical outcomes were considered exploratory.

A sample size of 8 patients per cohort (6 active, 2 placebo) was deemed appropriate to assess TD--0903 safety and tolerability during dose escalation. Safety, SaO2/FiO2 ratio, and efficacy data were summarised as descriptive statistics using SAS version 9.4 (SAS Institute, Cary, NC, USA).

Twenty-five patients enrolled and were randomised to receive TD-0903 1 mg (n = 6), 3 mg (n = 7), 10 mg (n = 6), or placebo (n = 6). One patient receiving TD-0903 3 mg was discontinued due to a negative SARS-CoV-2 PCR screening test returned after randomisation; per protocol, this subject was replaced.

Baseline data, concomitant medications, TEAEs, and clinical outcomes are summarised in the **Table**. Almost all patients (92%) received dexamethasone; 3 (12%) received remdesivir. The majority of TEAEs were mild to moderate and resolved by end of study, with no apparent dose relationship. Serious TEAEs occurred in 5 patients through Day 28, including COVID-19 progression in 1 placebo-treated patient; acute respiratory distress syndrome (ARDS) and fatal multiple organ dysfunction syndrome (MODS) in 1 placebo-treated patient; ARDS and fatal cardiac arrest in 1 placebo-treated patient; acute respiratory failure, ventricular fibrillation, and fatal MODS in 1 patient receiving TD-0903 1 mg; and ischaemic stroke in 1 patient receiving TD--0903 3 mg. No serious TEAEs were considered related to study treatment by the investigator. One patient receiving TD-0903 10 mg discontinued treatment on Day 4 due to an elevation in alanine aminotransferase (ALT) that resolved without consequence. No other changes in vital signs or laboratory safety measures, including creatinine and haematologic parameters, were attributed to treatment. The mean steady state maximal plasma concentration (C_max_) of TD-0903 at the highest dose was 19.0 ng/mL, well below levels predicted to produce systemic JAK inhibition.

**Table.**
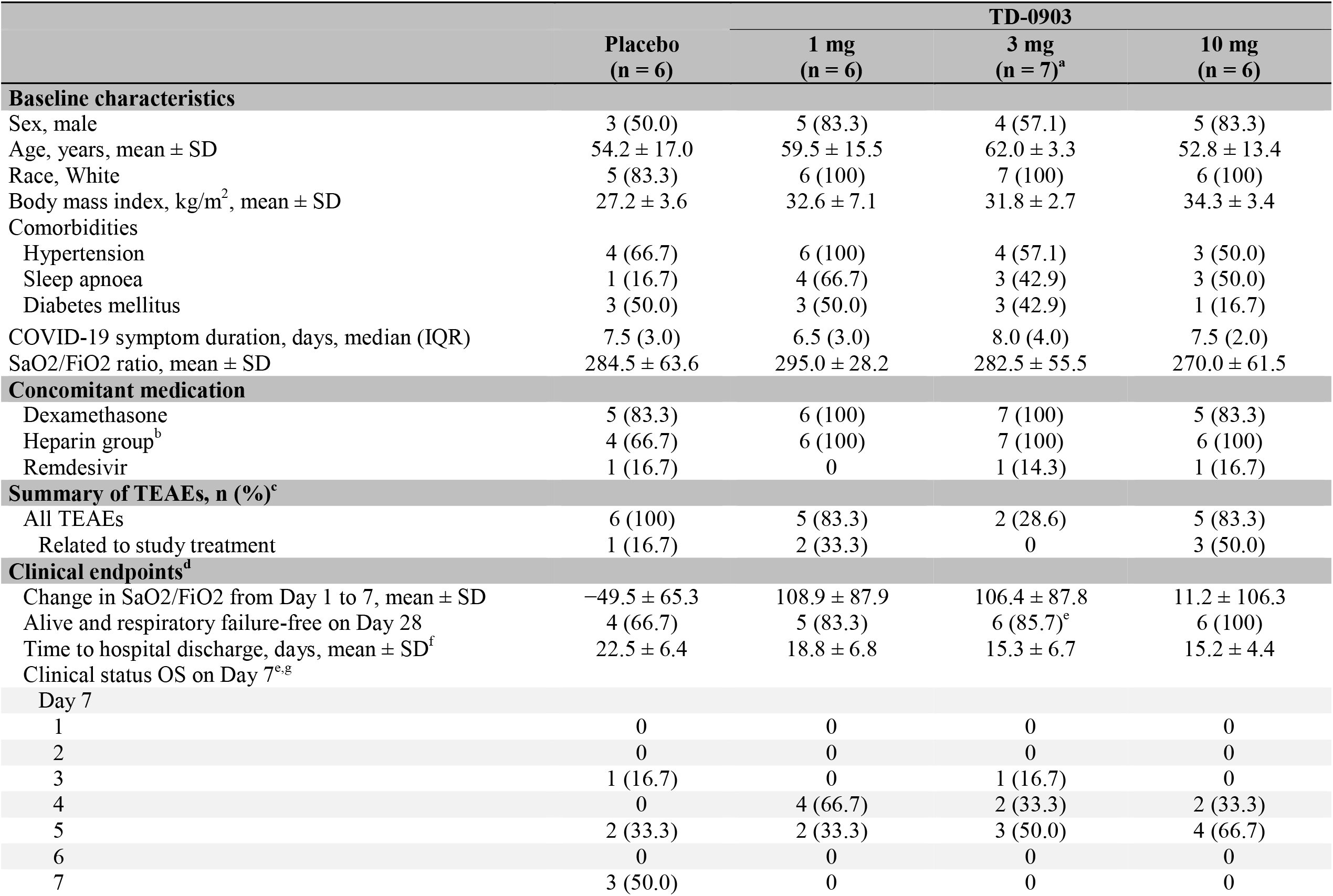
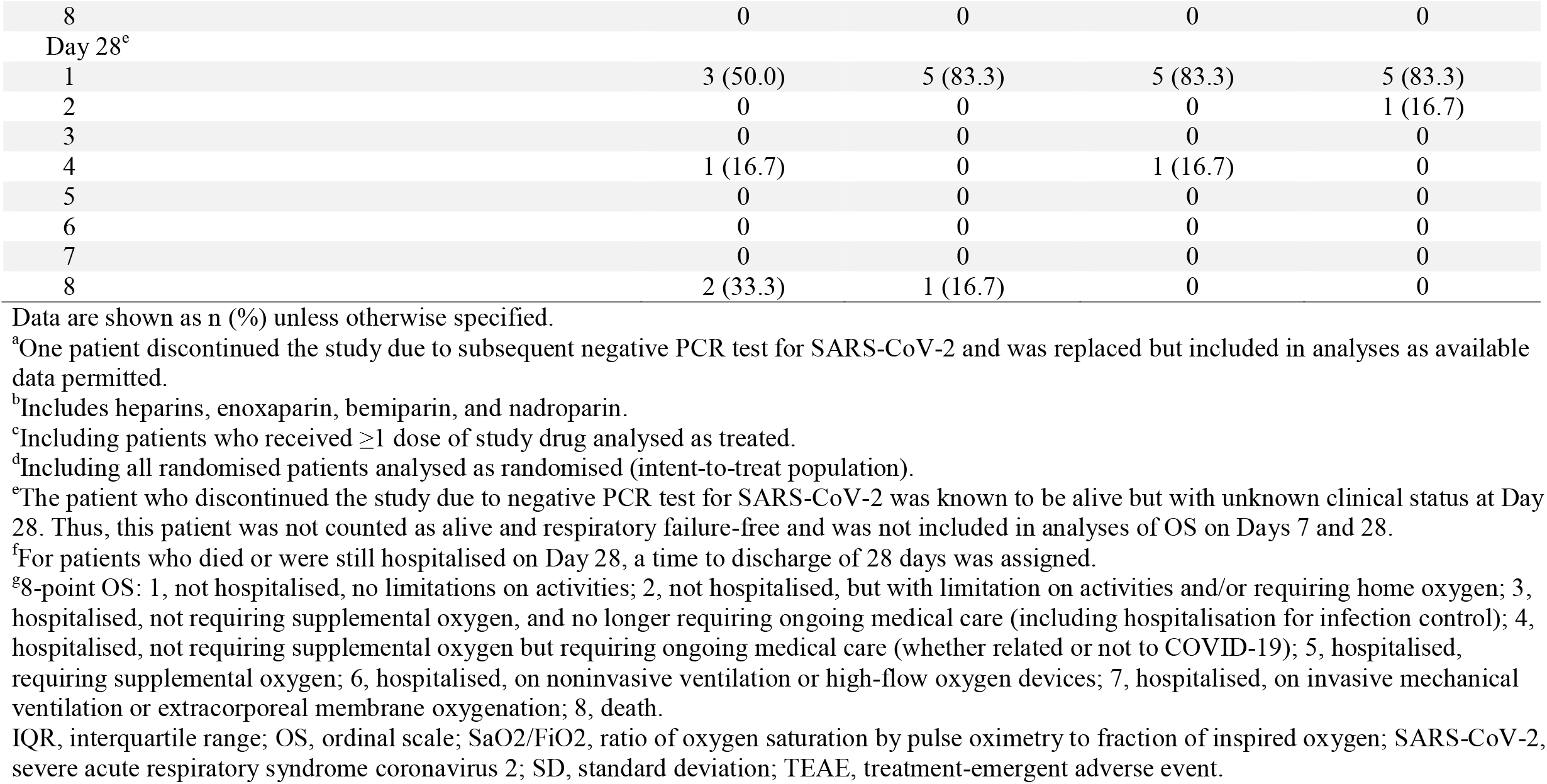
Key baseline data and outcomes

At baseline, all patients received supplemental oxygen via nasal prongs or mask (OS 5). Within the 7-day treatment period, 3 (50%) placebo-treated patients progressed to invasive mechanical ventilation (OS 7). In contrast, no patient receiving TD-0903 had a decline in clinical status. This pattern was maintained throughout the 28-day study period for patients receiving TD-0903 3 or 10 mg. One patient treated with TD-0903 1 mg and 2 placebo-treated patients died (OS 8) before Day 28 (Day 23 for patient treated with TD-0903 1 mg and Days 11 and 14 for placebo-treated patients); no patients treated with TD-0903 3 or 10 mg died during the 28-day study period. Improved SaO2/FiO2 ratio from baseline to Day 7 was observed in patients receiving TD-0903 vs placebo. The proportion of patients alive and respiratory failure-free at Day 28 was higher and mean time to hospital discharge was shorter in patients treated with TD-0903 vs placebo (**Table**).

This is the first clinical trial to date of delivery of a JAK inhibitor directly to the lung via inhalation in patients with COVID-19. Once-daily inhaled TD-0903 treatment for 7 days was generally well tolerated in patients with severe COVID-19. There were trends toward improvement in SaO2/FiO2 ratio, proportion of patients alive and respiratory failure-free at Day 28, and mean time to hospital discharge in patients treated with TD-0903 vs placebo. Overall mortality was 33% in placebo-treated patients vs 5% in patients treated with any dose of TD-0903. The small sample size of this early phase clinical trial limited evaluation of clinical efficacy through between-group comparisons and formal control for potential confounders. Nevertheless, the low mortality and pattern of earlier clinical recovery with TD-0903 vs placebo treatment suggests promise for the strategy of targeting cytokine-driven inflammation in the lungs of patients with severe COVID-19 through pan-JAK inhibition. Notably, JAK inhibition may have additive anti-inflammatory effects when used in combination with corticosteroid treatment[9]—which almost all patients in the study received—and thus using these treatments in combination may be complementary for the treatment of patients with COVID-19.

Based on the totality of evidence, inhaled TD-0903 3 mg was advanced for further evaluation in Part 2 of this study, a larger (N ≈ 200) double-blind, placebo-controlled parallel-group phase 2 study in hospitalised COVID-19 patients requiring supplemental oxygen (OS 5–6) (NCT04402866).

## Supporting information

CONSORT checklist

## Data Availability

Theravance Biopharma (and its affiliates) will not be sharing individual de-identified participant data or other relevant study documents.

## Acknowledgements

The authors and Theravance Biopharma, Inc., thank the patients and their families for their participation and Kyla Kennedy for clinical operations. Dave Singh, Robert Lord, and Ashley Woodcock are supported by the National Institute for Health Research (NIHR) Manchester Biomedical Research Centre (BRC).

## Contributors

**DS, EJM, RL, GDC, AW, BH**, and **RS** contributed to study design and interpretation. **DS, MB, VM**, and **RL** enrolled patients, collected data, and contributed to interpretation. **DLB** and **NDP** performed pharmacokinetic analyses and contributed to data interpretation. **JW** and **EK** contributed to data interpretation. **DAL, EYW**, and **TN** performed statistical analyses and contributed to data interpretation. All authors reviewed the manuscript critically for intellectual content and approved the final version for submission.

